# The Spectrum from Overt Primary Aldosteronism to Mild Dysregulated Aldosterone Production in Incidentally Discovered Adrenocortical Adenomas

**DOI:** 10.1101/2024.04.10.24305640

**Authors:** Thomas Uslar, Roberto Olmos, Alberth Burnier, Benjamín Sanfuentes, Pauline Böhm, Maria Paz Orellana, Francisco J. Guarda, Álvaro Huete, Nicolás Mertens, Cecilia Besa, Marcelo Andía, Alejandro Majerson, Jaime Cartes, Carlos Fardella, Fidel Allende, Sandra Solari, Anand Vaidya, Rene Baudrand

## Abstract

**Background:** Incidental adrenocortical adenomas (IA) are common. Current guidelines suggest screening for primary aldosteronism (PA) only in cases of hypertension or hypokalemia. This study aimed to evaluate the spectrum from overt PA to mild dysregulated aldosterone production with a sensitive protocol irrespective of blood pressure (BP) and potassium in patients with IA.

**Methods:** 254 consecutive patients (excluding hypercortisolism) were evaluated. The spectrum of PA was defined as a suppressed renin plus the following criteria: 1)Overt PA: aldosterone-to-renin-ratio (ARR) >30 ng/dL-to-ng/mL/hr, plasma aldosterone concentration (PAC) >15ng/dL, and/or 24h urinary aldosterone >10 ug/24h; 2)Moderate PA: ARR 20-30 ng/dL-to-ng/mL/hr, PAC 10-15 ng/dL; 3)Mild dysregulated aldosterone production: ARR <20 ng/dL-to-ng/mL/hr and PAC >5-10 ng/dL.

**Results:** 35% (n=89/254) met criteria for PA spectrum, 20% (34/89) were initially normotensive and 94% (84/89) normokalemic. Overt, moderate, and mild groups were 10%, 12%, and 13%. There were trends across groups of clinical severity: systolic BP (153±19, 140±14, 137±14 mmHg, p-trend<0.05), resistant hypertension (50%, 23%, 7% p-trend=<0.001), daily defined dose of antihypertensives (DDD) (3.2±1.6, 1.2±1.5, 0.4±0.6 p-trend=0.001), and lower eGFR (75.5±30.8, 97.8±38.5, 101±25.5, p-trend<0.01). At follow-up (mean 28±15 months), 87% had treatment with MR antagonists or surgery with decreased systolic BP relative to clinical severity, −31.3 ±23, −12.7 ±19, and −11.4 ±19 mmHg, (p-trend<0.001). Similar trends were observed for DDD, with significant increase in renin.

**Conclusions:** There is a prevalent spectrum of clinically-relevant PA and dysregulated aldosterone production in IA, irrespective of BP or potassium, usually undetected. Aldosterone-directed treatment improved BP and normalized renin even in milder cases.

**GRAPHIC ABSTRACT:** 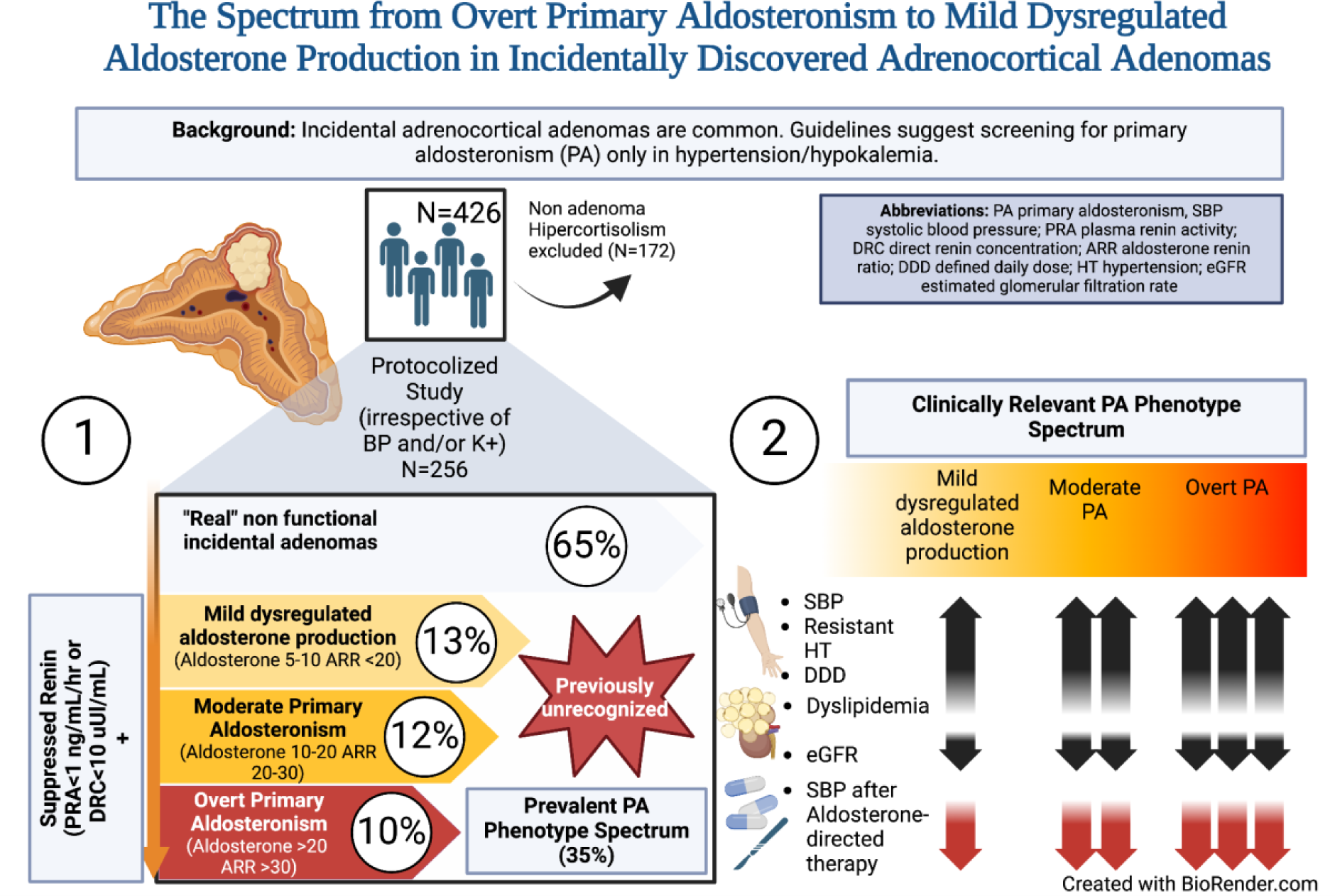

## INTRODUCTION

Incidental adrenocortical adenomas (IA) are increasingly identified with a prevalence of up to 6-10% in older adults.^1^ Clinical practice guidelines recommend screening all adrenocortical adenomas for cortisol excess. However, hormonal study for Primary Aldosteronism (PA) is currently advised only when patients with IA also have concomitant hypertension and/or hypokalemia, suggesting a screening selection bias.^1,2^

Over the last decades, a broader clinical phenotype of PA has been recognized. It is currently described that PA exists across a continuum of severity, from overt cases with advanced hypertension and/or hypokalemia, to milder forms that include individuals with prehypertension or even normotension.^3–5^ The clinical relevance of this continuum is highlighted in overt PA, a condition known to have substantially higher risk for adverse cardiovascular outcomes;^6,7^ however, it is increasingly recognized that patients with milder forms of PA also have a substantially higher risk for developing progressive cardiovascular disease, such as myocardial infarction, stroke, atrial fibrillation and left ventricular hypertrophy.^4,8–11^ Importantly, recognizing PA and implementing targeted therapy with mineralocorticoid receptor (MR) antagonists or surgery can mitigate these adverse cardiovascular outcomes in a real-life scenario.^6,12,13^

Taken together, this emerging evidence suggests that the current approach of PA screening in IA may be insensitive or incomplete.

We conducted a protocolized evaluation of consecutive patients with IA, irrespective of blood pressure status or serum potassium, to assess the PA phenotype spectrum and evaluate the clinical benefit of aldosterone-directed treatment during follow-up.

## METHODS

### Subjects

Patients with the diagnosis of adrenal incidentaloma as defined by the European Society of Endocrinology guidelines were recruited, treated, and followed at our Program for Adrenal Disorders between March 2017 and August 2023.^1,14^ Adenomas were defined by radiological assessment, as described below. Patients that met any of the following criteria were initially excluded: (i) age younger than 18 years, (ii) tumor size < 1 cm; (iii) known extra-adrenal metastatic disease; (iv) use of oral/intramuscular/intravenous glucocorticoids in the last 3 months or inhaled/transdermal glucocorticoids in the last month; and (v) clinically overt Cushing syndrome prior to adrenal radiological assessment.^14,15^ This study and our database was approved and monitored by the ethics committee of our institution. The participants gave written informed consent.

### Radiologic Assessment

Initially, a cohort of 426 consecutive subjects underwent an abdominal computed tomography (CT) scan, either nonenhanced or with iodinated intravenous contrast, in the setting of clinical conditions unrelated to adrenal disorders, where an adrenal incidentaloma was detected. The adrenal lesion was confirmed as an adenoma using attenuation criteria <10 Hounsfield units (HU) in unenhanced images or strict washout criteria when delayed contrast images were available (relative washout >40% and/or absolute washout >60%).^1,15^ When magnetic resonance imaging (MRI) was available, signal drop on out-of-phase images was considered diagnostic of an adenoma. When these methods were not available or diagnosis was unclear, confirmatory CT using a standard adrenal protocol was performed (noncontrast imaging, 70-second post-contrast venous phase imaging, and 10- to 15-minute delayed post-contrast phase), using known density and washout criteria for adenoma diagnosis.^1^ Applying these definitions, 83 subjects were excluded for having non-adenomatous adrenal masses (adrenocortical carcinoma, pheochromocytomas, metastasis, and other non-adenomatous lesions) (Fig. 1A).

**Figure 1.**
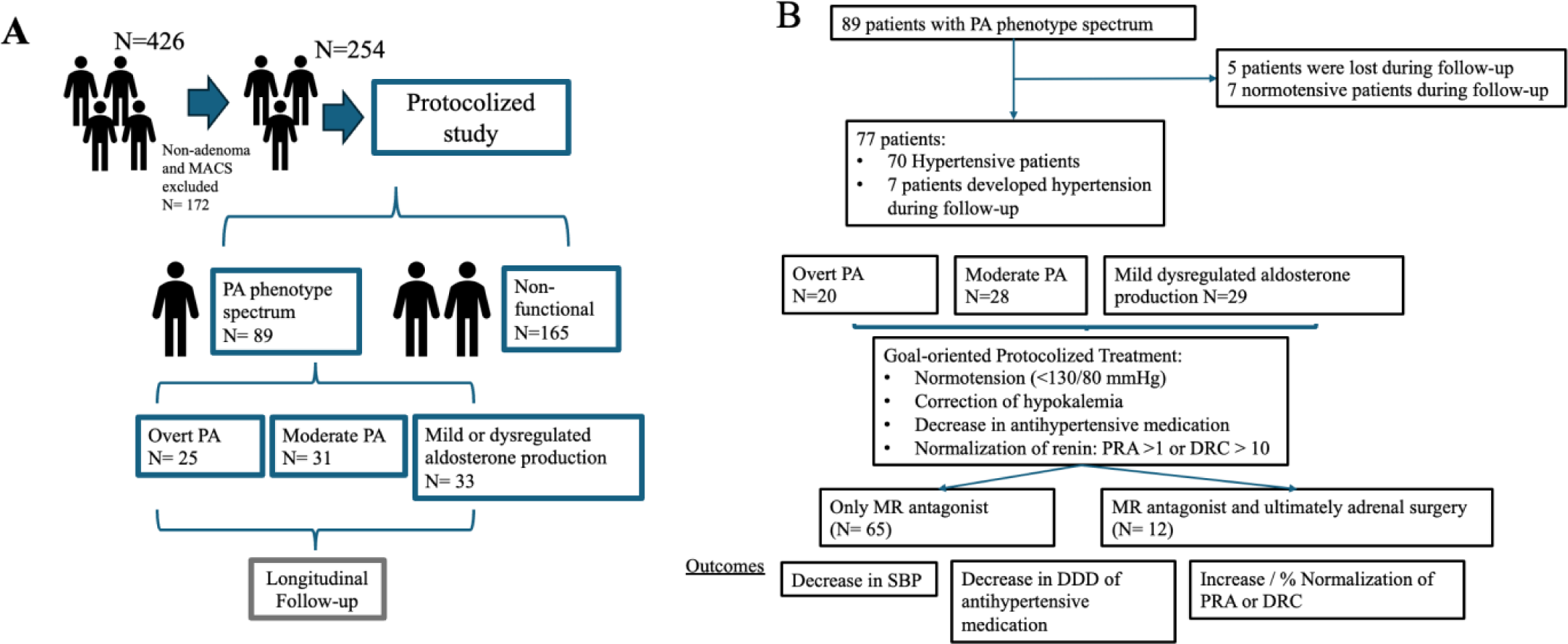
**A** Visual description of study design. **B** Derivation of longitudinal follow-up study cohort. MACS indicates mild autonomous cortisol secretion; PA, primary aldosteronism; DDD, defined daily dose; PRA, plasma renin activity; DRC, direct renin concentration.

### Endocrine Assessment

The Program for Adrenal Disorders at our institution has implemented a systematic hormonal assessment for all adrenal incidentalomas, as previously described.^14,15^ On the first day, blood samples were obtained between 08:00 and 09:00, after an 8-hour overnight fast, for measurement of seated plasma aldosterone concentration (PAC), plasma renin activity (PRA) and/or direct renin concentration (DRC), ACTH, serum creatinine, sodium and potassium. All patients submitted a 24h urine collection to measure urinary aldosterone, cortisol, cortisone and sodium. All subjects performed a 1-mg overnight dexamethasone suppression test (DST) wherein they took 1 mg of oral dexamethasone at 23:00, with measurement of serum cortisol on a subsequent day (between 08:00 and 09:00). In order to maximize sensitivity and avoid potential confounders, the samples were obtained under the following conditions: during follicular phase of the menstrual cycle in premenopausal women not taking estrogen-containing oral contraceptives; after at least two months without using estrogen-containing oral contraceptives or postmenopausal hormone replacement therapy (with the use of desogestrel if needed); patient in seated position after 30 minutes of rest; discontinuation of MR antagonist, potassium-sparing diuretics, and thiazides for at least one month before testing; avoidance of other interfering medications when used in moderate to high dose (angiotensin converting enzyme inhibitors, angiotensin II receptor blockers and beta-blockers) and use of non-interfering drugs such as amlodipine or doxazosin for blood pressure control during washout. All patients were advised to consume an unrestricted sodium diet 48h before screening to avoid false negative results as previously described.^2,15,16^

All blood samples were analyzed in the Clinical Laboratory of our institution, which meets international standards, such as the quality control program of the College of American Pathologist’s Laboratory Accreditation Program. Mild autonomous cortisol secretion (MACS) was defined as post-dexamethasone cortisol >1.8 ug/dL with a morning ACTH <15 pg/mL.^1^ Eighty-nine patients with adrenal adenomas (89/343: 26%) were diagnosed with MACS and were excluded from further analysis, reducing our final analyzed sample to 254 patients (Figure 1A).

The possibility of PA was clinically classified as a continuum of severity. To be classified as PA phenotype spectrum, all patients must have necessarily low or suppressed renin,^17,18^ defined as DRC <10 uIU/mL or PRA <1.0 ng/mL/hr.^19,20^ Among those who met the low renin criteria, the spectrum of PA was classified as: 1) Overt PA: aldosterone-to-renin ratio (ARR) >30 ng/dL-to-ng/mL/hr and either PAC >15 ng/dL and/or 24h urinary aldosterone >10 ug/24h; 2) Moderate PA: ARR 20-30 ng/dL-to-ng/mL/hr and PAC 10-15 ng/dL; 3) Mild dysregulated aldosterone production: ARR <20 ng/dL-to-ng/mL/hr and PAC >5-10 ng/dL.^20^ In order to use the same ARR threshold among the whole cohort, when DRC was measured, we divided DRC by 10, as previously described.^17,19,20^

### Longitudinal Follow-Up

IA patients with a PA phenotype spectrum were longitudinally assessed as shown in Figure 1B, but our analysis was retrospective, analyzing the impact of selected treatment. Once classified, patients with a PA phenotype spectrum who had hypertension received targeted aldosterone-directed treatment with either MR antagonist alone or MR antagonist and ultimately adrenal surgery decided by his/her physician. A goal-oriented, protocolized treatment was conducted, aiming for normotension (<120/80 mmHg), correction of hypokalemia if present pre-treatment, decrease in antihypertensive medication and normalization of renin (DRC >10 uIU/mL or PRA >1.0 ng/mL/hr) during follow-up. Whenever feasible, antihypertensive drug classes were simplified to include only MR antagonists. Since adrenal venous sampling is clinically unavailable in our country, the decision to pursue surgical adrenalectomy was evaluated by two experts using clinical criteria, including age, presence of hypokalemia, nodule size, aldosterone levels, and clinical/biochemical response to MR antagonist or adverse effects to medical treatment. Surgery was predominantly recommended for patients with unilateral nodules, young individuals (age <40-50 years) presenting with hypokalemia, and PAC >20 ng/dL, as described in our previous report.^21^ Regarding treatment with MR antagonists, spironolactone was predominantly prescribed for female patients, while eplerenone was the preferred choice for male patients. Normotensive patients at enrollment were monitored during follow-up without treatment, however, if they developed incident hypertension during follow-up, aldosterone-directed therapy was initiated. Following the initiation of targeted therapy, follow-up visits were conducted wherein patients with a PA phenotype spectrum were reassessed after a mean of 28±15 months. Five patients did not attend follow-up appointments and were excluded for further analysis (Figure 1B). A complete set of follow-up data, including the same laboratory protocol as described above, was analyzed on those who met criteria for PA phenotype spectrum.

### Hormonal Assays

PAC and DRC were determined by chemiluminescence analyzer (LIAISON®XL; DiaSorin); intra-assay coefficient of variation (CV) was <9.5% and <13% respectively. Serum cortisol was determined by electrochemiluminescence immunoassay (ECLIA, Cobas; Roche); intra-assay CV was <5%. ACTH was determined by chemiluminescent immunoenzymatic solid-phase assay (Immulite®2000 XPi; Siemens); intra-assay CV was <10%. Urinary aldosterone, cortisol and cortisone were measured by LC-MS/MS as previously described.^22^ Urinary cortisol/cortisone ratio was used as a proxy of 11HSDB2 dysfunction, a well-known cause of low renin hypertension.^23^

### Comorbidities

All participants underwent physical examination [body weight (kilograms), height (centimeters), and body mass index (BMI; kilograms per square meter)] and ambulatory measurements of systolic and diastolic blood pressure (SBP and DBP, respectively) with an authorized automatized arm BP device.^24^ Hypertension was defined as a SBP of at least 130 mm Hg and/or a DBP of at least 80 mm Hg measured at least twice, on different days.^24^ Resistant hypertension was defined as above-goal elevated blood pressure (BP) in a patient despite the concurrent use of 3 antihypertensive drug classes, including a diuretic. Resistant hypertension also includes patients whose BP achieves target values on ≥4 antihypertensive medications.^25^ Antihypertensive medication was quantified as a defined daily dose (DDD), which is the assumed average maintenance dose per day for a drug used for its main indication in adults.^26^ Dysglycemia (prediabetes or diabetes) was defined by documented diagnosis and/or a hemoglobin A1c >5.7% or a serum glucose >140 mg/dL after a 2-hour oral glucose tolerance test.^27^ Dyslipidemia was defined by calculated LDL-cholesterol >130 mg/dL, triglycerides >150 mg/dL or HDL-cholesterol <40 mg/dL in males or <50 mg/dL in females, or use of proper medication.^28^ Estimated glomerular filtration rate (eGFR) was calculated with the Cockroft-Gault equation.^29^

### Statistical Analysis

Continuous variables are presented as mean and SD. Categorical comparisons were performed with Chi-square test and continuous variables were compared using Student t-test, with P value of <0.05 considered significant. Normality was assessed by Kolmogorov–Smirnov test. When variables were not normally distributed, a bootstrapping analysis with resampling in 1000 iterations was applied to deal with outliers and nonnormality issues, and to confirm the internal validation of the analysis. Furthermore, a linear regression model was performed with unadjusted and adjusted models (by age and sex) to assess the association of the 3 phenotypes of PA spectrum with clinical parameters. Adjusted linear regression and ANOVA analyzing the phenotypes of PA spectrum were also performed. Paired T-tests were used to assess the treatment outcomes at follow-up. Statistical analyses and graphics were performed using SPSS software (v.28:Inc., Chicago, IL) and Prism10.

## RESULTS

### Study Population and Demographics

The demographic data and comorbidities of the 254 consecutive patients with IA and no evidence of hypercortisolism are presented in Table 1. Left-sided adenomas were more common than right-sided adenomas and the average size was 19±8.5 mm. There was a high rate of hypertension (53%) and resistant hypertension (13%) as well as other metabolic disorders.

**Table 1.** Baseline characteristics for the 254 patients with incidental adrenocortical adenomas, after excluding hypercortisolism.

|  | Incidental adrenocortical adenomas without hypercortisolism (N=254) |
| --- | --- |
| Age (yr) | 55±12 (range 18-87) |
| Female (%) | 77 |
| BMI (Kg/m<sup>2</sup>) | 28.6±4 |
| Hypertension at baseline (%) | 53 |
| Resistant hypertension (%) | 13 |
| DDD of antihypertensive medication | 1.04 (2) |
| Dyslipidemia (%) | 47 |
| Dysglycemia (%) | 53 |
| Adenoma size (mm) | 19±8.5 |
| Left-side (%) | 54 |
| Plasma aldosterone (ng/dL) | 12.3±15.8 |
| Suppressed renin (DRC <10 uIU/mL or PRA <1 ng/mL/hr, %) | 47% |
| ARR (ng/dL)/(ng/mL/h) | 21±60 |
| Urinary aldosterone (ug/24h) | 7.1±6.5 |
| Plasma creatinine | 0.77±0.2 |
| Plasma potassium | 4.2±0.4 |

Values are mean±SD. DDD= Defined daily dose, value is mean (IQR)
Abbreviations: BMI, body mass index; DDD, defined daily dose; DRC, direct renin concentration; PRA, plasma renin activity; ARR, aldosterone-to-renin ratio.

### Comparing the PA phenotype spectrum versus non-functionality in IA

Notably, 35% (89/254) of patients with IA had low renin levels and met at least one criterion for having a PA phenotype spectrum, as shown in Table 2. Of note, 20% (18/89) of these individuals were initially normotensive at recruitment and 94% (84/89) were normokalemic. When compared to individuals with non-functioning IA (n=165/254), patients with a PA phenotype spectrum had a higher prevalence of hypertension (80 vs 48% p<0.05), resistant hypertension (22 vs 7% p<0.05) and dyslipidemia (63 vs 36% p<0.001). There was a trend towards a higher prevalence of hypokalemia in patients within the PA phenotype spectrum; however, this trend did not reach statistical significance. No differences were observed by groups in dysglycemia, urinary sodium excretion or urinary cortisol/cortisone ratio (proxy of 11BHSD2 activity), which are well known modulators of low renin levels.

**Table 2.** Clinical characteristics of patients with and without primary aldosteronism (PA) phenotype spectrum.

|  | All PA phenotype spectrum (N=89, 35%) | Non-functioning adenoma (N=165, 65%) | P value |
| --- | --- | --- | --- |
| Age (yrs) | 57±11 | 54±12 | 0.618 |
| Female (%) | 75 | 79 | 0.438 |
| **Hypertension (%)** | **80** | **48** | **<0.001** |
| **Resistant hypertension (%)** | **22** | **7** | **0.020** |
| Hypokalemia (%) | 6 | 2 | 0.119 |
| **Dyslipidemia (%)** | **63** | **36** | **<0.001** |
| Obesity (%) | 41 | 34 | 0.373 |
| Dysglycemia (%) | 50 | 54 | 0.609 |
| Urinary sodium (mEq/24h) | 187 | 168 | 0.705 |
| Urinary F/E ratio | 0.32 | 0.31 | 0.759 |

Abbreviations: PA, primary aldosteronism; F/E, cortisol/cortisone ratio

### The severity spectrum of the PA phenotype in IA

Overt PA, moderate PA, and mild dysregulated aldosterone production were identified in 10%, 12%, and 13% of the overall sample, respectively, as shown in Table 3.

**Table 3.**
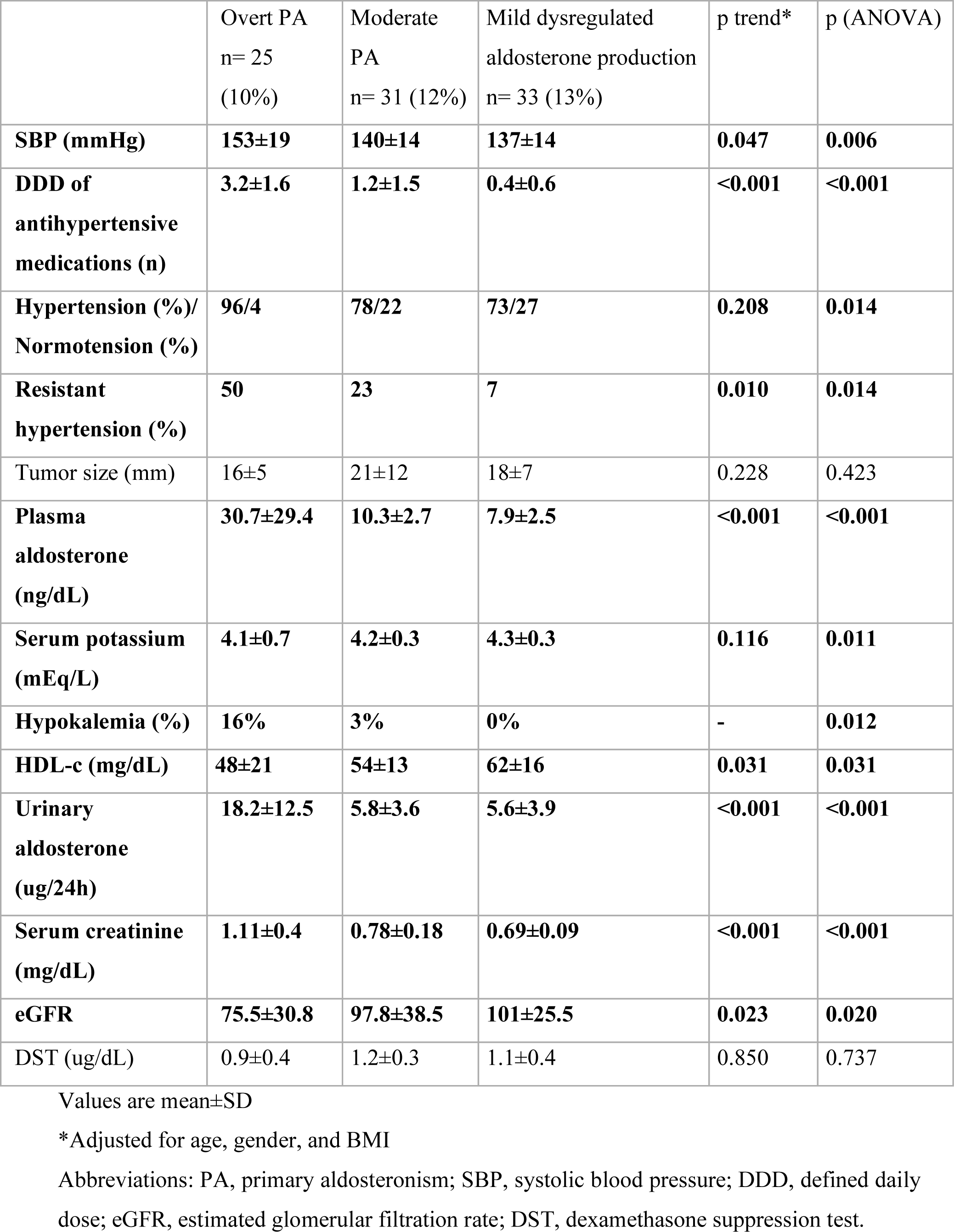
Analysis of the clinical impact of progressive phenotypes of primary aldosteronism spectrum.

|  | Overt PA<br>n= 25<br>(10%) | Moderate<br>PA<br>n= 31 (12%) | Mild dysregulated<br>aldosterone production<br>n= 33 (13%) | p trend* | p (ANOVA) |
| --- | --- | --- | --- | --- | --- |
| **SBP (mmHg)** | **153±19** | **140±14** | **137±14** | **0.047** | **0.006** |
| **DDD of**<br>**antihypertensive**<br>**medications (n)** | **3.2±1.6** | **1.2±1.5** | **0.4±0.6** | **<0.001** | **<0.001** |
| **Hypertension (%)/**<br>**Normotension (%)** | **96/4** | **78/22** | **73/27** | **0.208** | **0.014** |
| **Resistant**<br>**hypertension (%)** | **50** | **23** | **7** | **0.010** | **0.014** |
| Tumor size (mm) | 16±5 | 21±12 | 18±7 | 0.228 | 0.423 |
| **Plasma**<br>**aldosterone**<br>**(ng/dL)** | **30.7±29.4** | **10.3±2.7** | **7.9±2.5** | **<0.001** | **<0.001** |
| **Serum potassium**<br>**(mEq/L)** | **4.1±0.7** | **4.2±0.3** | **4.3±0.3** | **0.116** | **0.011** |
| **Hypokalemia (%)** | **16%** | **3%** | **0%** | **-** | **0.012** |
| **HDL-c (mg/dL)** | **48±21** | **54±13** | **62±16** | **0.031** | **0.031** |
| **Urinary**<br>**aldosterone**<br>**(ug/24h)** | **18.2±12.5** | **5.8±3.6** | **5.6±3.9** | **<0.001** | **<0.001** |
| **Serum creatinine**<br>**(mg/dL)** | **1.11±0.4** | **0.78±0.18** | **0.69±0.09** | **<0.001** | **<0.001** |
| **eGFR** | **75.5±30.8** | **97.8±38.5** | **101±25.5** | **0.023** | **0.020** |
| DST (ug/dL) | 0.9±0.4 | 1.2±0.3 | 1.1±0.4 | 0.850 | 0.737 |

Values are mean±SD
\*Adjusted for age, gender, and BMI
Abbreviations: PA, primary aldosteronism; SBP, systolic blood pressure; DDD, defined daily dose; eGFR, estimated glomerular filtration rate; DST, dexamethasone suppression test.

Notably, when comparing these 3 groups, there were parallel trends in the severity of SBP, prevalence of resistant hypertension, DDD of antihypertensive medication, magnitude of hypokalemia and plasma and urinary aldosterone production. Moreover, the severity of the PA phenotype spectrum was associated with lower HDL-cholesterol, higher serum creatinine and lower estimated glomerular filtration rate.

Also, there were no differences in tumor size, serum potassium and DST between PA phenotypes.

### Treatment Follow-Up

The mean length of follow-up was 28±15 months. During this time, 50% (n=7/14) of initially normotensive individuals developed hypertension.

The distribution of new-onset hypertension cases in the severity spectrum of PA was the following: one patient with overt PA, 2 patients with moderate PA, and 4 patients with mild dysregulated aldosterone production. Eighty-seven percent (n=77/89) of patients with hypertension and PA spectrum phenotype who received aldosterone-directed treatment had complete data after longitudinal follow-up, of which 84% (n=65/77) were treated only with MR antagonist and 16% (n=12/77) were treated with MR antagonist and ultimately unilateral adrenalectomy (Figure 1B). We observed a clinical benefit of targeted aldosterone-directed therapy which paralleled the severity of the PA phenotype (Table 4); patients with overt PA had the greatest reductions in SBP and DDD of antihypertensive medication with aldosterone-directed therapy, with milder PA phenotypes displaying lesser reductions. Notably, all patients exhibited a significant increase in renin after treatment follow-up, irrespective of the treatment modality or their group categorization within the PA spectrum.

**Table 4.**
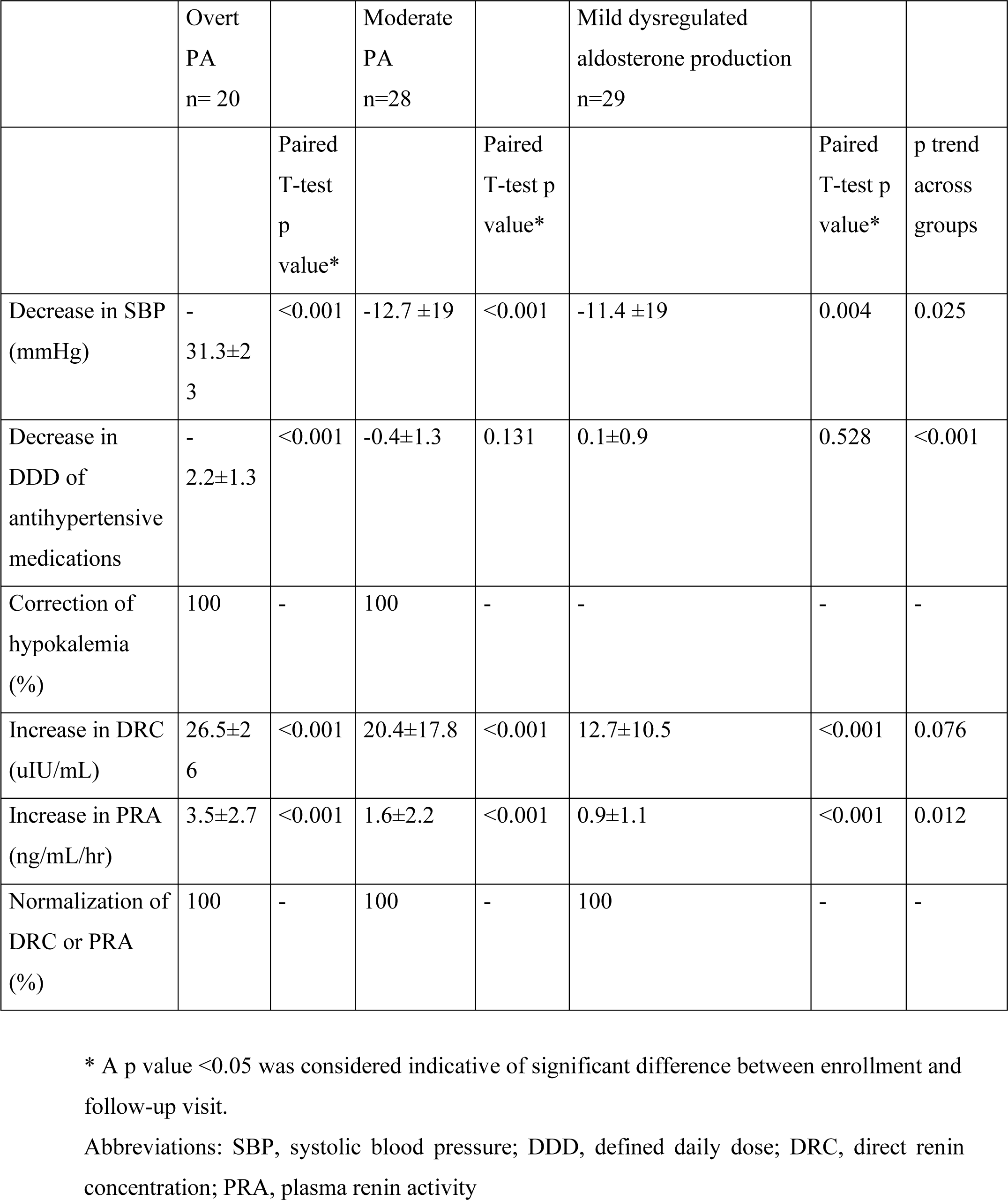
Analysis of response to aldosterone-directed treatment after follow-up in patients with primary aldosteronism (PA) phenotype spectrum.

|  | Overt<br>PA<br>n= 20 |  | Moderate<br>PA<br>n=28 |  | Mild dysregulated<br>aldosterone production<br>n=29 |  |  |
| --- | --- | --- | --- | --- | --- | --- | --- |
|  |  | Paired<br>T-test<br>p<br>value* |  | Paired<br>T-test p<br>value* |  | Paired<br>T-test p<br>value* | p trend<br>across<br>groups |
| Decrease in SBP<br>(mmHg) | -<br>31.3±2<br>3 | <0.001 | -12.7 ±19 | <0.001 | -11.4 ±19 | 0.004 | 0.025 |
| Decrease in<br>DDD of<br>antihypertensive<br>medications | -<br>2.2±1.3 | <0.001 | -0.4±1.3 | 0.131 | 0.1±0.9 | 0.528 | <0.001 |
| Correction of<br>hypokalemia<br>(%) | 100 | - | 100 | - | - | - | - |
| Increase in DRC<br>(uIU/mL) | 26.5±2<br>6 | <0.001 | 20.4±17.8 | <0.001 | 12.7±10.5 | <0.001 | 0.076 |
| Increase in PRA<br>(ng/mL/hr) | 3.5±2.7 | <0.001 | 1.6±2.2 | <0.001 | 0.9±1.1 | <0.001 | 0.012 |
| Normalization of<br>DRC or PRA<br>(%) | 100 | - | 100 | - | 100 | - | - |

\* A p value <0.05 was considered indicative of significant difference between enrollment and follow-up visit.
Abbreviations: SBP, systolic blood pressure; DDD, defined daily dose; DRC, direct renin concentration; PRA, plasma renin activity

## DISCUSSION

### Summary of Main Findings

In this protocolized study, we evaluated consecutive patients with IA to show that overt PA, as well as a continuum of milder phenotypes of dysregulated aldosterone production, can be frequently detected in up to one third of IA cases, even among patients with normal blood pressure and serum potassium.

The clinical relevance of this spectrum of PA in patients with IA is highlighted when targeted therapy with MR antagonists or surgery resulted in improvements in BP and antihypertensive medication profiles after raising renin levels, even in mild cases. Considering that clinical practice guidelines on IA emphasize screening for PA only in the setting of hypertension and/or hypokalemia, our findings support an expanded screening for PA irrespective of BP or potassium, since most milder cases of aldosterone dysregulation may currently be misdiagnosed as “non-functioning”.

### Comparison to Previous Studies

Historically, most adrenocortical adenomas have been suspected to be non-functioning and, therefore, hormonal analysis is not usually performed in non-endocrinological settings (only 10% of IA in some studies, only 30% in others). Thus, functional status is clearly being underdiagnosed.^30,31^ However, other authors have recently described a much higher incidence of aldosterone autonomy than previously reported. In a prospective study with adrenal incidentaloma patients, Kmieć et al. showed higher rates of autonomous aldosterone secretion than classic cohorts by using more sensitive criteria of ARR >10 (ng/dL)/(ng/mL/h)) (approximately) and unsuppressed PAC >4 ng/dL.^32^ Overall, 25.4% of individuals with adrenal incidentaloma were found to have dysregulated aldosterone production. Consistently, we found that our permissive criteria for PA spectrum was associated with higher frequency of hypertension, resistant hypertension, dyslipidemia and lower eGFR when compared to “real” non-functioning adrenocortical adenomas.

There is biological plausibility in our results. A spectrum of MR activation from low renin hypertension to overt PA has been described, with aldosterone producing cell clusters and age as main modulators of zona glomerulosa activity.^5,33–35^

It is possible that adrenocortical adenomas may not always be the source of aldosterone autonomy in some milder subjects, but the presence of a nodule may trigger screening and reveal an abnormal baseline adrenal dysregulated aldosterone production.^36^ This is consistent with the histological finding that CYP11B2-positive areas are not uncommon in patients with PA and adenomas, even when cross-sectional imaging detects no radiological abnormalities in normal adrenal tissue.^34,36^

### Clinical implications

The data from this study clearly showed various degrees of renin-independent aldosterone production in patients who would have otherwise been labeled as having “non-functioning” IA, despite having higher cardiovascular morbimortality, as previously described.^37^ Those studies did not systematically assess PA phenotype spectrum, therefore, at least part of this excess cardiovascular morbimortality might be attributed to aldosterone dysregulation. Consistently, we demonstrated higher rates of hypertension, resistant hypertension, dyslipidemia and lower eGFR in PA phenotype spectrum compared to real non-functioning IA.

Another implication of our findings is that patients with IA, suppressed renin and “inappropriate” normal plasma aldosterone concentration, may require careful assessment depending on the clinical real-life scenario: (1) Hypertensive patients, even with mild dysregulated aldosterone production, could benefit from receiving aldosterone-directed treatment with MR antagonists or surgery, as we observed a significant reduction in SBP, and a better hypertensive profile (lower or same number of antihypertensive medication but better chosen in the context of low renin hypertension) while observing a raise in renin levels in all cases, proportionate to severity. (2) Normotensive patients with PA phenotype spectrum should be closely monitored, as we found that 50% of them developed arterial hypertension after short term follow-up. This is concordant with our report in which patients with normotensive mild PA had an increased risk for hypertension by probable increased MR activity.^4^

As expected, the impact of targeted aldosterone directed therapy in reducing both SBP and antihypertensives was more pronounced with the increasing severity of PA phenotype spectrum, but was not limited only to overt cases, maintaining clinical benefit in SBP even in mild dysregulated aldosterone production. The DDD of antihypertensive drugs significantly decreased in patients with overt PA, but not in those with moderate or mild dysregulated aldosterone production. This finding could be attributed to the fact that these patients initially had lower baseline BP, requiring either no or few antihypertensive medications. Additionally, the high prevalence of newly diagnosed arterial hypertension among patients in those groups may have contributed to this finding.^5^

### Limitations and Strengths

Our study has some limitations. Our longitudinal data is restricted to almost 3 years, and we did not fully measure end-organ damage and major cardiovascular consequences of dysregulated aldosterone production. Nevertheless, clear improvements in renin and blood pressure were observed with targeted aldosterone-directed therapy, which are well-known proxies for better cardiovascular outcomes. This was achieved even without the use of confirmatory testing, which might have reduced the number of patients who could potentially benefit from targeted therapy due to stricter thresholds.^3,38,39^ Additionally, PAC was measured using immunoassay, which has known limitations compared to LC-MS/MS,^40^ however, urinary aldosterone was measured using LC-MS/MS, and the results behaved similarly.

Ultimately, the absence of adrenal vein sampling (AVS) in our country may have inclined us towards medical rather than surgical treatment for more patients. However, a recent study has suggested that AVS cannot reliably exclude asymmetric bilateral aldosterone secretion in patients with aldosterone producing adenomas,^41^ and our treatment approach still yielded better SBP, reduced medication usage and improved renin levels.

The strengths of our study include a structured and meticulous protocol performed in all subjects without selection bias. Also, we used permissive criteria to accurately phenotype hormone production through multiple measurements, as well as implementing careful targeted therapy for MR activation in selected cases.

## PERSPECTIVES

Our protocolized study shows that there is a prevalent spectrum of clinically-relevant PA and dysregulated aldosterone production among individuals with IA, irrespective of blood pressure or potassium levels. Due to probable bias in screening for primary aldosteronism (PA) in patients with IA based on blood pressure and cumbersome diagnostic methods, milder cases of aldosterone dysregulation often go undetected by current clinical approaches, leading to inadequate labeling as ‘non-functioning’. Our follow-up data confirmed that treatment with MR antagonists or surgery in a real-life scenario improved BP, decreased the number of medications and normalized renin levels even in milder cases, outcomes that are not usually achieved by current hypertensive medications that do not directly decrease MR activation. As the field of PA phenotype spectrum is rapidly evolving, further studies are warranted to explore the long-term cardiovascular outcomes of targeted therapy to decrease MR activation in patients with mild dysregulated aldosterone production in IA.

## NOVELTY AND RELEVANCE

- What is new? In patients with IA, unbiased screening for PA regardless of hypertension status or hypokalemia improved the diagnosis of prevalent and clinically relevant disease spectrum. Follow-up data showed benefits of aldosterone-targeted treatment even in milder cases.
- What is relevant? Our results highlight the importance of improved screening for a PA spectrum of disease among patients with IA, since many of them are currently misdiagnosed as “non-functional” and are not offered proper treatment, despite having higher cardiovascular morbidity.
- Clinical implications?
- If we improve the detection of PA spectrum in patients with IA, we can offer readily-available specific therapies that may reduce cardiovascular morbimortality in future studies.

## Data Availability

Data is available if requested and will be provided under restrictions to protect confidential persona information of patients.

## Article Information

### Author Contributions

R.Baudrand designed the study. T.Uslar, R.Olmos, A.Burnier, B.Sanfuentes, P.Böhm and FJ.Guarda collected samples and clinical data from patients. A.Huete, C.Besa, N.Mertens, M.Andia performed radiological studies. J.Cartes and A.Majerson performed adrenal surgeries. F.Allende and S.Solari performed laboratory studies. T.Uslar performed statistical analysis and wrote the first draft of the article. All authors contributed to writing the article and approved the final version to be published. T.Uslar and R.Baudrand had full access to all the data in the study and had final responsibility for the decision to submit for publication.

## Acknowledgments

We are thankful to the adrenal clinical care and research team.

## Non-standard Abbreviations and Acronyms

IA: incidental adrenocortical adenomas
PA: primary aldosteronism
MR: mineralocorticoid receptor
CT: computed tomography
HU: Hounsfield units
MRI: Magnetic resonance imaging
PAC: plasma aldosterone concentration
PRA: plasma renin activity
DRC: direct renin concentration
DST: dexamethasone suppression test
MACS: mild autonomous cortisol secretion
ARR: aldosterone-to-renin ratio
DDD: defined daily dose.

## Supplemental Material

**Figure S1.**
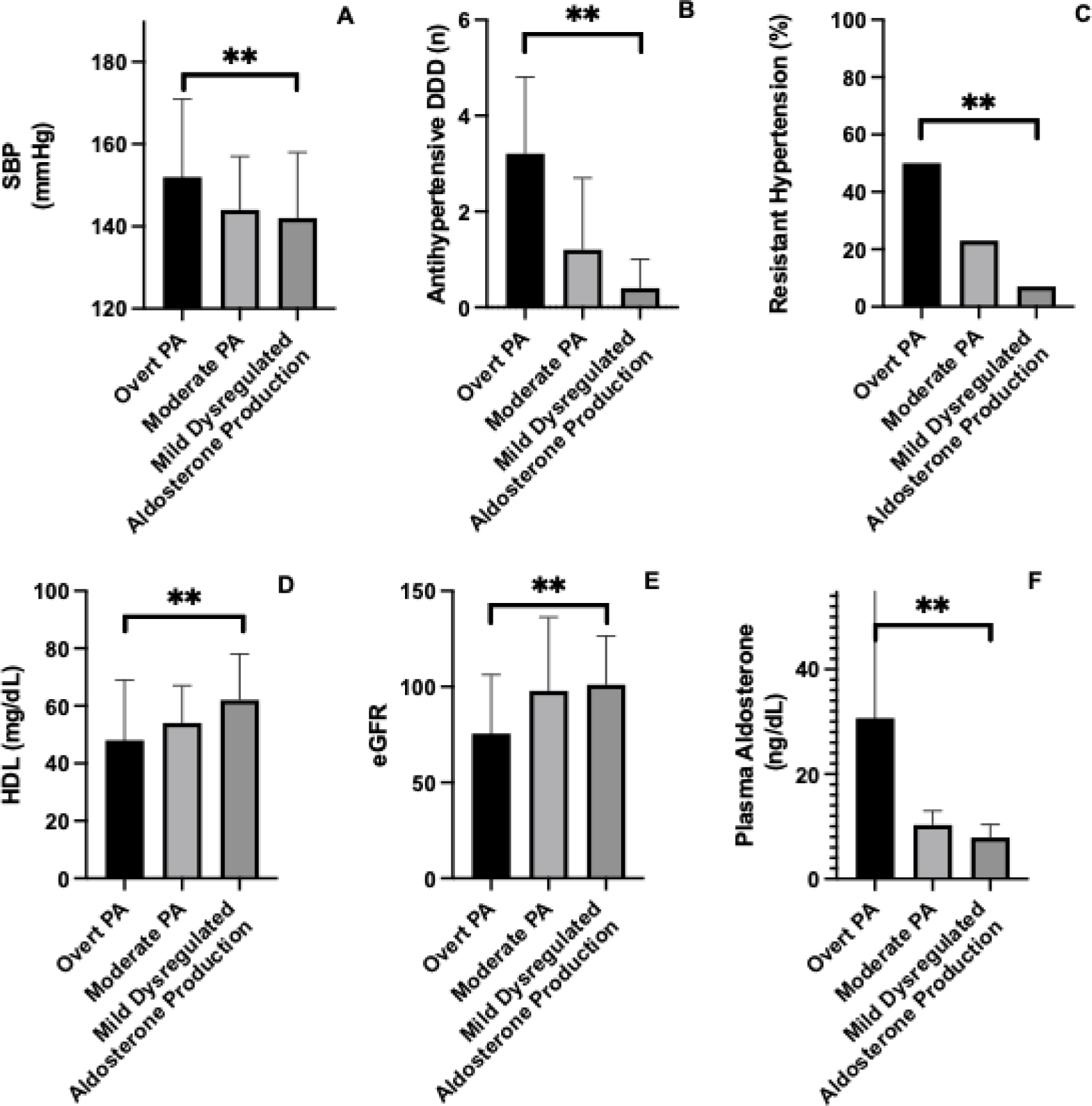
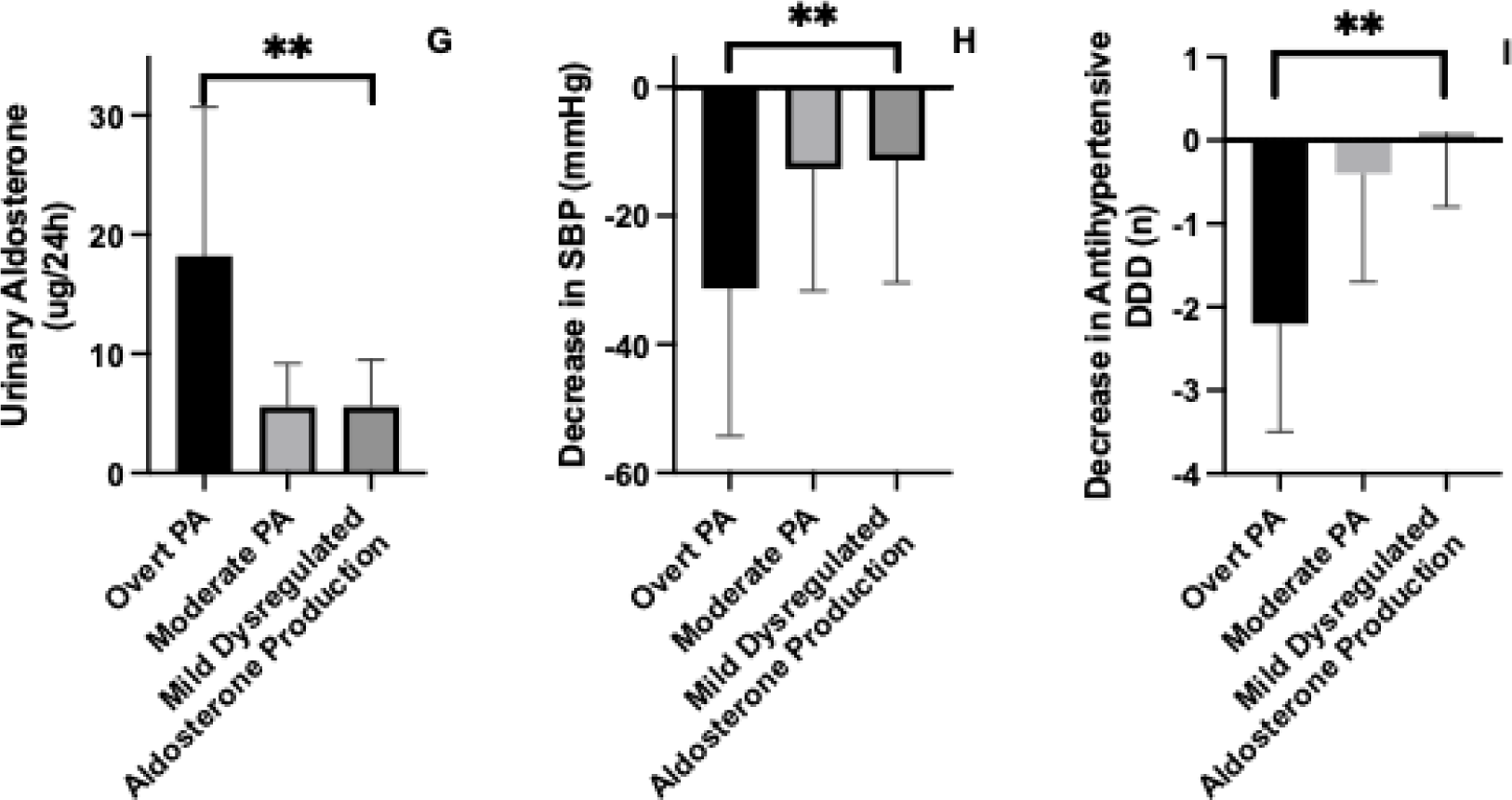
Analysis of the clinical impact of primary aldosteronism phenotype spectrum. (A) SBP across spectrum of primary aldosteronism groups, (B) DDD of antihypertensive medications, (C) rate of resistant hypertension, (D) HDL-cholesterol levels, (E) eGFR, (F) plasma aldosterone, (G) urinary aldosterone, (H) Decrease of SBP after treatment, (I) Decrease of DDD of antihypertensive medications after treatment. The boxplots show the mean and SD of each group. Abbreviations: SBP, systolic blood pressure; DDD, defined daily dose; eGFR estimated glomerular filtration rate; SD, standard deviation.

